# Safety and Effectiveness of Iota-Carrageenan Eye Drops for the Treatment of Dry Eye Disease

**DOI:** 10.64898/2026.03.16.26348126

**Authors:** Marta Blanco Vázquez, Margarita Calonge, Hanna Dellago, Nicole Unger-Manhart, Markus Savli, Sabine Roch-Nakowitsch, Eva Prieschl-Grassauer

## Abstract

**Purpose:** The aim of this clinical investigation was to evaluate the safety and effectiveness of iota-carrageenan (I-C) eye drops in the treatment of mild-to-moderate dry eye disease (DED).

**Methods:** In this prospective, single arm, open label clinical investigation, thirty adult participants with mild-to-moderate DED applied I-C eye drops three times daily for four weeks. Before start and after end of treatment, participants rated DED symptoms (foreign body sensation, burning/stinging, itching, pain, sticky feeling, blurred vision and photophobia), both after exposure to a normal controlled environment (NCE) and to an adverse controlled environment (ACE). Additional endpoints were changes in ocular surface disease index (OSDI), Change in Dry Eye Symptoms Questionnaire (CDES-Q), corneal and conjunctival surface damage, tear film break-up time, tear evaporation and production. Tolerability was assessed by participants at start and end of treatment. Safety, including visual acuity and intraocular pressure, was monitored throughout the investigation.

**Results:** After four weeks of treatment with I-C eye drops, the mean total DED score after ACE was significantly reduced by −11.89 points (95% CI: −15.11, −8.67), p<0.001. The mean score reduction between baseline and final visit after NCE was slightly less pronounced, with −8.07 points (95% CI: −10.71; −5.43), p<0.001. The vast majority of participants (93% after ACE and 89% after NCE exposure) recorded a reduction in total DED score between baseline and final visit. Mean OSDI score significantly decreased by −7.75 points (95% CI: −10.85, −4.63), p<0.001. ACE-induced deterioration of tear film stability as well as corneal and conjunctival damage were reduced following treatment. All adverse events were mild and transient in nature. 93% of the patients described I-C eye drops as well or very well tolerated. Treatment did not negatively impact any of the safety parameters.

**Conclusion:** I-C eye drops are effective, safe and well tolerated. Treatment with I-C eye drops alleviates DED symptoms, stabilizes tear film and protects the ocular surface in patients with mild-to-moderate DED even under adverse environmental conditions.

**Trial Registration:** NCT06262100.

## Introduction

Dry eye disease (DED), also termed dry eye syndrome (DES), is a multifactorial disease of the ocular surface characterized by a loss of homeostasis of the tear film and accompanied by ocular symptoms. Common symptoms of DED are burning or stinging sensations, foreign body sensation, grittiness, light sensitivity, fluctuant, blurred or impaired vision, and mucus secretion. Symptom severity reaches from discomfort to disabling ocular pain and can severely affect vision-related quality of life by limiting activities such as working, driving, reading, as well as sport and other recreational activities.^1^

Tear-film instability and hyperosmotic stress, ocular surface inflammation and damage, as well as neurosensory abnormalities play etiological roles. DED can occur on its own or as a consequence of other underlying diseases like Sjögren’s syndrome, diabetes, thyroid eye disease, or blepharitis. Among the risk factors for developing DED are environmental, sociodemographic and lifestyle factors like climate conditions, age, allergies, smoking, contact lens use, excessive electronic screen use, intake of certain medications (eg, topical glaucoma medication containing benzalkonium chloride (BAK), antihistamines, antidepressants and antianxiety medications, beta blockers, diuretics and oral corticosteroids), as well as surgical procedures to the eye. Women are more prone to develop DED than men.^1,2^ The global prevalence of DED ranges from 5% to 50%, depending on the respective geographic region and age group.^3^

To restore and maintain a healthy ocular surface, cornea and conjunctiva must be kept permanently wet and lubricated. Ideally this function is fulfilled by the tear film. Therefore, DED therapy aims at adequate lubrication of the ocular surface by restoring homeostasis of the tear film.^4–6^ Mild to moderate cases are usually treated with a range of lubricating ophthalmic solutions, gels and ointments that can be easily purchased over-the-counter (OTC). While a variety of ophthalmic solutions based, eg, on polyvinyl alcohol, glycerin or cellulose derivatives like hydroxypropylmethylcellulose (HPMC) and carboxymethylcellulose (CMC),^4^ the market in the last decades has been dominated by products based on hyaluronic acid (HA). Besides the introduction of perfluorohexyloctane eye drops,^7,8^ innovation in the DED therapeutic area has mainly taken the form of new combinations of existing compounds, primarily HA.

Given the heterogeneous nature of the disease and the limitations of current treatments, like tolerability issues, limited ability to promote regeneration or healing of ocular tissue damage, or delay of several weeks or months in the onset of therapeutic effect,^9^ there is a growing demand for new therapeutic strategies that widen the variety of available treatment options and address the considerable unmet needs of the market.^10^

Here, we propose iota-carrageenan (I-C) eye drops as a promising, safe and effective alternative for the treatment of dry eyes. Carrageenans are natural polysaccharides built of D-galactose and 3, 6-anhydro-D-galactose units. They are obtained by extraction of certain red seaweeds of the Rhodophyceae class. In the food, cosmetic and pharmaceutic industry, carrageenans are widely utilized due to their excellent physical functional properties, such as gelling, thickening, emulsifying and stabilizing abilities.^11^ I-C holds great promise for use in lubricating and moisturizing eye drops based on the following properties: It forms an in situ gelling system upon exposure to physiological conditions,^12^ and hence retains water on the ocular surface. It increases viscosity of a solution which enhances its retention time on the ocular surface. In addition, it is a non-Newtonian polymer and as such it is per definition shear-thinning, meaning that the viscosity decreases with an increasing shear rate, thus reducing resistance against movements of the eyelid when blinking.^13^ Finally, I-C acts locally and is not absorbed by the body, therefore it may be used also during pregnancy and lactation.

The suitability of I-C for ophthalmic applications has already been demonstrated in various contexts: Carrageenans have been tested and proposed as promising carriers for ocular drug delivery.^14–16^ A commercial I-C-containing viscous gel has been proven to be safe and effective when used to protect and hydrate the corneal surface during ophthalmic surgery.^17,18^ We have previously demonstrated the hydrating, lubricant capacity of iota-carrageenan and its ability to protect the ocular surface from dehydration in a cell culture model and in an ex vivo animal model (unpublished data). Consequently, we have developed I-C eye drops that were placed on the market in compliance with the medical device directive 93/42/EEC. The present clinical investigation is proposed to evaluate the lubricating performance of I-C eye drops when treating patients diagnosed with mild-to-moderate DED. Results are intended to demonstrate conformity with the new medical device regulation (MDR) (2017/745).

## Material and methods

### Study Design

The clinical investigation presented in this paper aimed to evaluate the safety and performance of I-C eye drops in the treatment of mild-to-moderate DED. This was a non-randomized, uncontrolled, open-label, single arm, monocentric study. The investigational product (IP) is CE-marked under the Medical Device Directive (MDD). The study is registered on clinicaltrials.gov under the identifier NCT06262100.

After the screening visit (visit 1, day −7), participants underwent a 7-day washout period with three times daily use of saline eye drops (Hydrabak ^®^, Thea S.A.) until the day before the next visit (visit 2, baseline assessment, day 0 ± 2). A follow-up visit (visit 3) and the final visit (visit 4) were scheduled on day 14 ± 2 and day 28 ± 4, respectively. At visits 2 and 4, participants were first exposed to a normal controlled environment (NCE, 23°C, 50% relative humidity, no airflow) for 30 minutes, followed by exposure to an adverse controlled environment (ACE, 23°C, 10% relative humidity, airflow) for 90 minutes. Ophthalmologic examinations were conducted after NCE and after ACE. The first application of the IP was administered by the investigator at visit 2 (baseline visit), after the baseline ophthalmologic examinations. Participants were provided with the IP and trained in self-administration. They were instructed to administer one drop of the IP per eye three times per day (early morning, midday, and bedtime) for 28 ± 4 days. At visit 3, only NCE (not ACE) exposure and evaluation were conducted, while participants continued self-administration of the IP for another two weeks. Participants were instructed to administer the last dose of IP 2 ± 1 hour before attending the final visit (visit 4), where NCE and ACE evaluation and compliance check were done. The total study duration for the individual participant was 35 ± 6 days.

Throughout the study, participants were asked to document IP administration, adverse events and use of concomitant medication in the patient diary.

Figure 1 shows the graphical abstract of the clinical study, outlining the timeline of visits and treatments.

**Figure 1.**
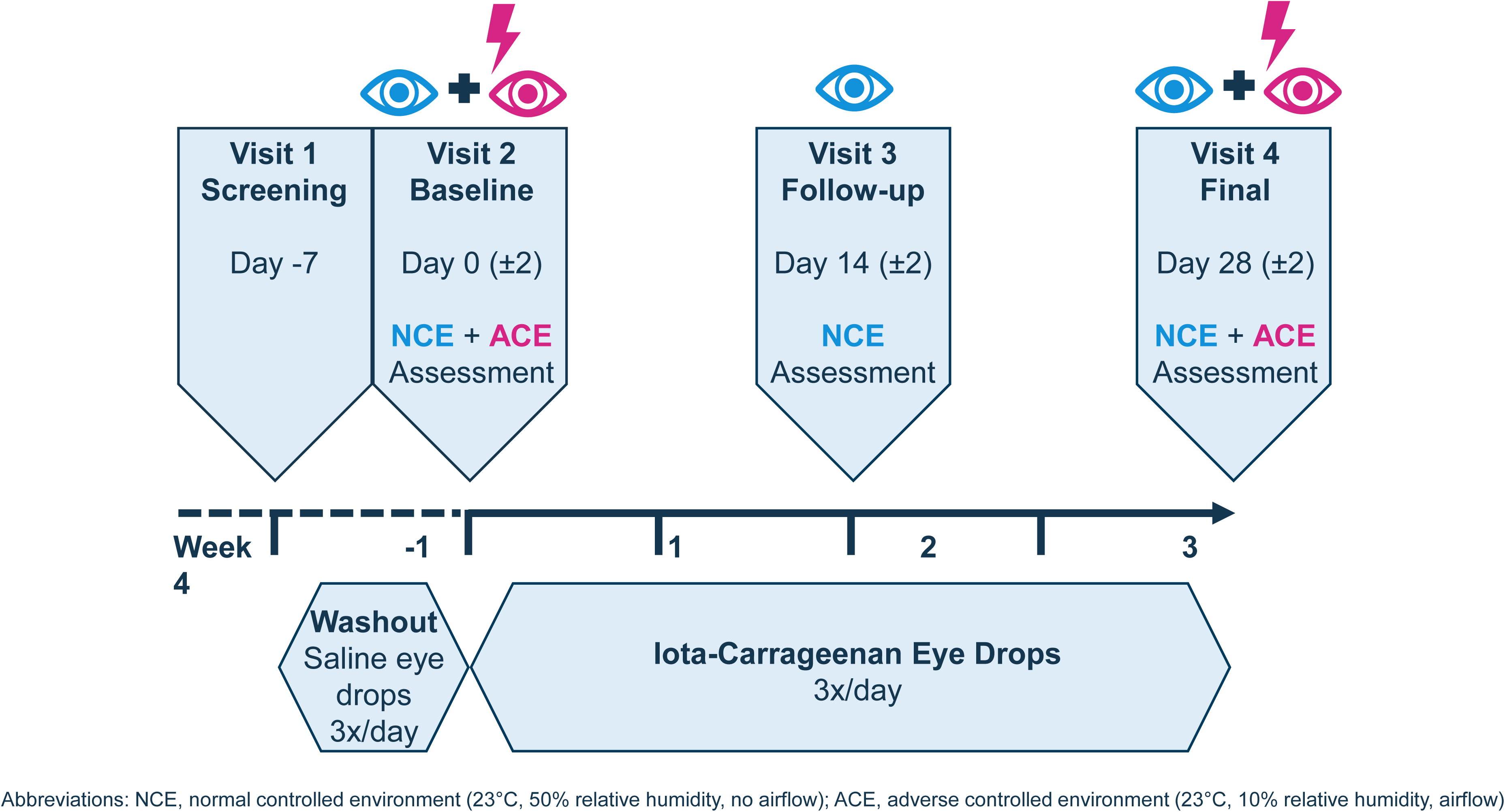
Study Overview.

### Participants

Study participants were adults of any sex who had been diagnosed by an ophthalmologist with mild-to-moderate DED for at least three months before study start. Mild-to-moderate DED was defined as an Ocular Surface Disease Index (OSDI) score ≥ 13 and < 33 points. The following DED seriousness criteria precluded participation in the study: fluorescein corneal staining score ≥ 4 in the Oxford scale; lissamine green conjunctival staining score ≥ 4 in the Oxford scale; Schirmer test score < 2 mm; fluorescein TBUT = 0 seconds. To be included in the study, participants had to provide written informed consent, be willing and able to administer the study eye drops themselves in both eyes and abstain from use of contact lenses and of any other topical ocular medication for the intended duration of the study.

Exclusion criteria were 1) known hypersensitivity, allergy, or intolerance to the IP or any of its components; 2) any ocular surface disease or condition other than DED; 3) any other active ocular inflammatory disease or condition, glaucoma or any ocular surgery during the last 6 months; 4) lacrimal punctum occlusion during the previous month or planned during the study period; 5) any other topical ocular medication, such as non-steroidal anti-inflammatory drugs, etc. within 30 days of the inclusion visit (visit 2); 6) clinically relevant or uncontrolled systemic diseases (except for controlled Sjögren syndrome) during the last 3 months that may interfere with study assessments or results; 7) topical ocular treatment with calcineurin inhibitors (such as cyclosporin or tacrolimus) during the last 12 weeks, corticosteroids during the last four weeks, blood derivatives, insulin or amniotic membrane preparations during the last week, or during the last month before the screening visit (visit 1); 8) treatment with any ocular lubricant after the screening visit (visit 1) other than the saline eyedrops (Hydrabak^®^); and 9) systemic treatments that may affect DED signs or symptoms, ocular surface or vision.

Women of childbearing potential had to commit to using effective methods of contraception during the entire study period and have a negative pregnancy test result at the screening visit.

### Interventions and Procedures

Participants administered one drop of the IP per eye three times per day for 28 ± 4 days. The IP, iota-carrageenan (I-C) eye drops, is a preservative-free sterile buffered ophthalmic solution containing 3.2 mg/ml I-C. The IP was manufactured and labeled according to Good Manufacturing Practice (GMP) by Hälsa Pharma GmbH, Lübeck, Germany.

This study included both exposure to standard conditions (normal controlled environment, NCE) to assess baseline symptoms, as well as to adverse conditions (adverse controlled environment, ACE) to elicit DED symptoms following the protocols of the Controlled Environment Laboratory (CELab), Vision R&D at the Institute of Applied Ophthalmobiology (IOBA) of the University of Valladolid, Spain. Some ophthalmologic examinations were performed after exposure to NCE conditions, some after ACE, and some, including the primary effectiveness endpoint, were evaluated under both conditions. Table 1 provides an overview of procedures and examinations conducted at each visit, following NCE and ACE, listed in temporal order from the least to most invasive. Since DED severity can fluctuate throughout the day, all assessments were conducted at the same time of day across all visits for each participant.

**Table 1.** Ophthalmologic examinations carried out at each visit.

| Visit number | Visit 1 | Visit 2 |  | Visit 3 | Visit 4 |  |
| --- | --- | --- | --- | --- | --- | --- |
| Description | Screening | Baseline |  | Follow-up | Final |  |
| Timing | Day -7 | Day 0 (±2) |  | Day 14 (±2) | Day 28 (±4) |  |
| After type of environment | n.a. | NCE | ACE | NCE | NCE | ACE |
| Total DED symptom score (prim. endpoint) | x | x | x | x | x | x |
| OSDI questionnaire | x | x |  |  | x |  |
| CDES-Q |  |  |  |  | X<sup>a</sup> |  |
| BCVA (ETDRS) | x | x |  | x | x |  |
| NIBUT | x | x | x |  | x | x |
| IOP |  | x |  | x | x |  |
| Slit-lamp examination | x | x | x | x | x | x |
| TBUT | x | x | x |  | x | x |
| Corneal fluorescein staining | x | x | x | x | x | x |
| Conjunctival lissamine green staining | x | x | x |  | x | x |
| Schirmer test without anaesthesia | x |  | x |  |  | x |

<sup>a</sup> Assessed with respect to the status at the Baseline visit.
**Abbreviations:** ACE, adverse controlled environment (for 90 minutes); BCVA, best corrected visual acuity;
CDES-Q, Change in Dry Eye Symptoms Questionnaire; DED, dry eye disease; ETDRS, Early Treatment of
Diabetic Retinopathy Study; IOP, intraocular pressure; NCE, normal controlled environment (for 30 minutes);
NIBUT, non-invasive tear breakup time; OSDI, Ocular Surface Disease Index; TBUT, tear breakup time.

Tolerability was assessed on a Likert-type scale. Safety was assessed by systematic recording of adverse events (AE) occurrence, severity, seriousness, causal relationship to the IP, and outcome. Visual acuity, intraocular pressure and slit lamp examination of the ocular surface and anterior intraocular structures were additional safety endpoints.

Please refer to the Supplemental Methods for a detailed description of the effectiveness and safety assessments.

### Endpoints

The primary effectiveness endpoint of the clinical investigation was the total score of DED related ocular symptoms collected through numerical rating scales (NRS). The total DED score was determined by adding up the ratings for seven symptoms: foreign body sensation, burning or stinging, itching, pain, sticky feeling, blurred vision, and photophobia, with each symptom being self-assessed by the participants on a scale of 0 to 10, resulting in a maximum total score of 70 points.

Analysis of the primary endpoint was based on the mean total DED score difference between baseline and final visit after ACE exposure. Mean total DED score difference after NCE exposure were analyzed as a secondary endpoint. The key secondary effectiveness endpoint was the patient responder rate evaluated as per the total DED score.

Additional secondary effectiveness endpoints were:

- Score difference on OSDI questionnaire.
- Evaluation of Change in Dry Eye Symptoms Questionnaire (CDES Q).
- Corneal fluorescein staining pattern assessed with the Corneal and Contact Lens Research Unit (CCLRU) grading scale and the Oxford grading scale.
- Conjunctival lissamine green staining pattern assessed with the Oxford scale.
- Tear film evaporation assessed by evaporimetry.
- Stability of tear film assessed by Tear Break-Up Time (TBUT) and non-invasive TBUT (NIBUT).
- Tear production according to the Schirmer test without anaesthesia.

The safety endpoints were:

- Best corrected visual acuity (BCVA) evaluated as per the Early Treatment Diabetic Retinopathy Study (ETDRS) procedure.
- Slit lamp examination of the ocular surface and anterior intraocular structures;
- Intraocular pressure.
- Tolerability level of the IP as evaluated by participants by means of a Likert-type scale;
- Occurrence of adverse events (AEs).

### Sample Size Calculation

The primary variable for calculating the necessary sample size was the total DED score, which can range from 0 to 70.

The null hypothesis (H0) was defined as: Following the usage of the IP, the difference between the baseline and the final measurement of the total DED score is 0.

The alternative hypothesis (H1) was defined as: The difference between the baseline and the final measurement of the total DED score is not equal to 0.

Expected values necessary for sample size calculation were derived from a published clinical study.^19^ In that study, a mean difference from baseline to day 21 of 195.6 points (standard deviation [SD] 84.4) was reported for participants treated with hyaluronic acid, based on the visual analogue scale (VAS), which is similar to the DED score, but with a range of 0 to 700. Considering this data and the transformation from a 0-700 scale to a 0-70 scale, we have estimated the expected mean difference of the total DED score to be around 7 points in a more conservative scenario (less than half of the mean difference observed in the cited study).

Additionally, an increased SD of 12 points was expected. To achieve a statistical power of 80% at a two-sided alpha level of 0.05, while assuming an effect size of 0.58 (7/12), a sample size of 26 participants was required. Considering a potential dropout rate of up to 15%, the total necessary sample size was 30 participants.

### Statistical Analysis

The final data analysis was performed after the finalization and approval of the SAP and after database lock. Effectiveness was analyzed in the full analysis set (FAS) and the per-protocol set (PPS). The FAS comprised all participants to whom the IP has been administered and was regarded as the primary effectiveness analysis population. Since effectiveness analyses were based on comparisons between baseline and post-treatment, results were analyzed in participants with data available for all visits, ie, the evaluable FAS. The PPS comprised all participants in the FAS with no major protocol violation and with self-administration compliance of at least 80%. The difference was tested for statistical significance through the paired t-test. A significant effectiveness of the IP, ie, a significant difference between total DED scores at baseline and final visit, was to be declared if the 95% confidence interval (CI) for the score difference did not encompass zero. The mean score difference between baseline and final visit after NCE exposure was analyzed in an analogous manner. Responder rate (number of participants with a reduction in the total DED score) was analyzed for both NCE and ACE.

For secondary endpoints OSDI score, corneal and conjunctival tear film evaporation, tear break-up time (NIBUT and TBUT), and tear production, comparisons were made between baseline and final visits. P values were derived by paired t-test or by bootstrapping. The analysis of the CDES-Q was conducted descriptively. Additional secondary endpoints include the corneal fluorescein staining pattern assessed using the CCLRU and Oxford grading scales, and the conjunctival lissamine green staining pattern assessed using the Oxford grading scale. Comparisons were made between baseline and final visits, separately for NCE and ACE. For the Oxford Score, the average of left and right eye was used to calculate mean values. Although Oxford grading is *per se* ordinal, several published studies have treated aggregated corneal staining scores as quasi-continuous, as has been done here for descriptive purposes. ^20–22^

In general, secondary endpoints were analyzed in an explorative sense. Statistical tests and the p-values attached to them should be regarded as descriptive and not as tests of hypotheses.

Safety endpoints were analyzed in the safety population, which comprised all enrolled participants that complied with the inclusion/exclusion criteria and have received at least one dose of the IP. AEs were coded based on the Medical Dictionary for Regulatory Activities (MedDRA) and summarized descriptively by system organ class (SOC) and preferred term (PT), as well as by seriousness, severity, causal relationship to the IP, action taken, and outcome.

When calculating the incidence of AEs (i.e., on a per-participant basis), each participant was counted once, and any repetitions of AEs within a participant were ignored for events coded in the same category.

R version 4.0.5 was used to generate tables, figures, and listings and statistical analyses, unless otherwise noted.

## Results

### Participant Disposition and Baseline Characteristics

The clinical investigation was conducted between December 2023 and June 2024. A total of 30 participants with mild to moderate DED were screened after giving informed consent. All 30 screened volunteers complied with all inclusion and exclusion criteria and were enrolled, thus constituting the safety population and the FAS. Two participants terminated their participation in the clinical investigation prematurely: one due to an unrelated AE and one due to poor tolerance of the IP. Two participants achieved less than the predefined 80% compliance for IP self-administration. Hence, 26 participants completed the clinical investigation as per protocol. Figure 2 shows a flow chart detailing participant allocation and analysis sets.

**Figure 2.**
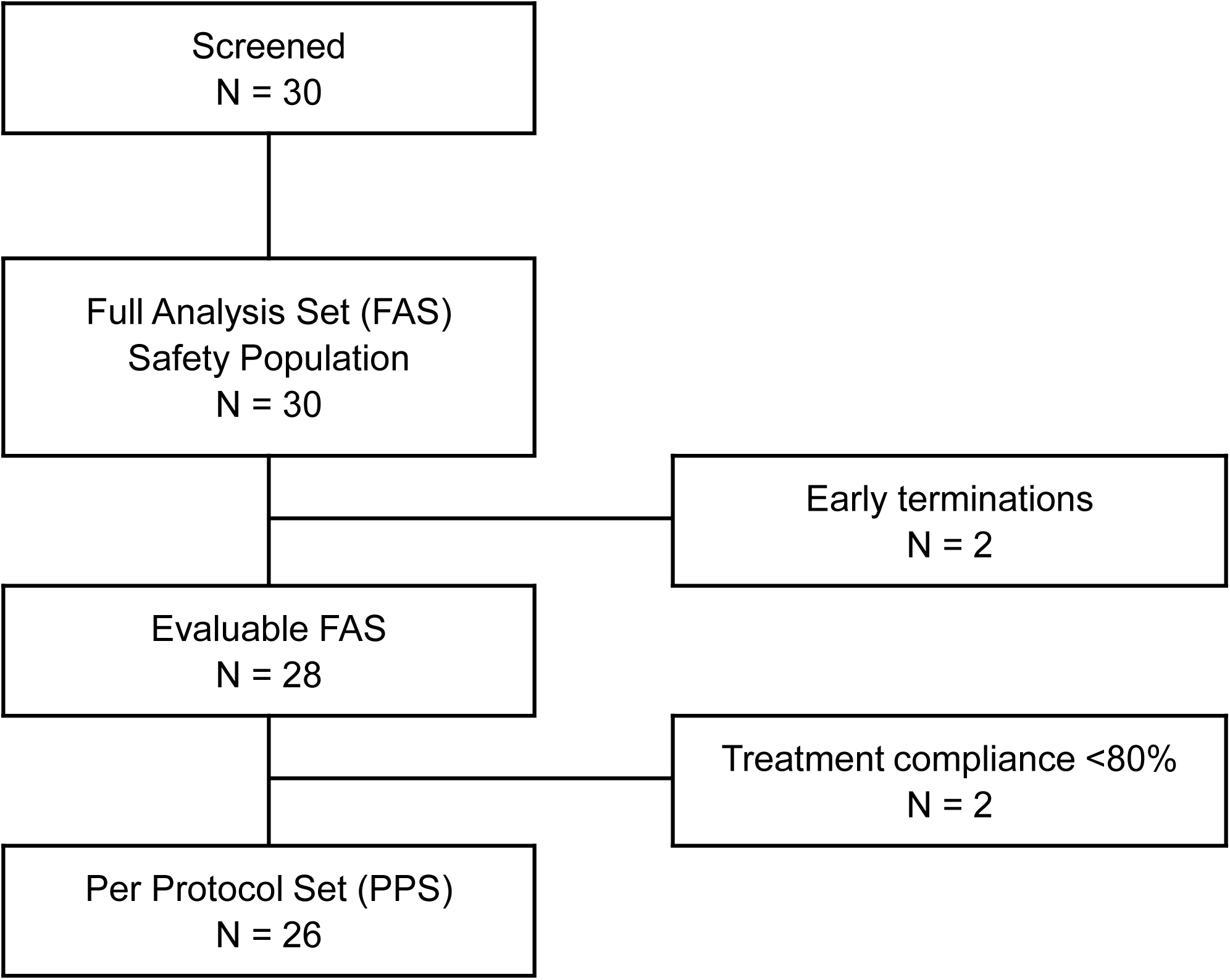
Flow chart depicting flow of participants through the study.

Demographic characteristics are summarized in Table 2. Overall, 20 (67%) of the 30 participants were female. Mean age was 50.6 (SD 14.33, range 22-77) years. 93% were caucasian and 7% were black.

**Table 2.** Summary of demographic and baseline characteristics, safety population.

| Parameter | Statistic | Total<br>N=30 |
| --- | --- | --- |
| Sex - n (%) | Female | 20 (66.67) |
|  | Male | 10 (33.33) |
| Ethnic origin - n (%) | White | 28 (93.33) |
|  | Black | 2 (6.67) |
|  | Other | 0 (0) |
| Age (years) | Mean (SD) | 50.63 (14.33) |
|  | Median | 57.50 |
|  | Q1, Q3 | 40.25, 61.00 |
|  | Min, Max | 22, 77 |

**Abbreviations:** N, number of participants in safety population; n, number of participants in respective category; SD, standard deviation; Q1, first quartile; Q3, third quartile; min, minimum value; max, maximum value.

### Effectiveness

All effectiveness results are presented for the evaluable FAS, ie, participants in the FAS with data available for all visits. Results for the PPS were similar to those for the FAS, ie, only minor numerical differences were observed between FAS and PPS.

The primary endpoint was the change in DED symptom intensity between baseline and final visit, ie, after four weeks of three times daily treatment with the IP. For the primary endpoint analysis, the change in ocular symptoms was assessed based on the total DED score rated by participants after ACE exposure, and for secondary endpoint analysis after NCE exposure. After ACE exposure, the mean (SD) DED score was reduced from 21.89 (11.07) at baseline to 10.00 (8.78) at the final visit. This corresponds to a 54% reduction in symptom severity, indicating marked improvement in DED symptoms after ACE over the four-week treatment period. The mean difference between baseline and final visit after ACE exposure was −11.89 (95% CI −15.11; −8.67), with a p value for the difference of < 0.001 (paired t-test). Based on the 95% CI boundaries not encompassing the zero value, the null hypothesis was rejected and a significant effect of the IP on DED symptoms can be declared.

After NCE exposure, the mean DED score difference between baseline and final visit was −8.07 (95% CI −10.71; −5.43) points, again with a p value for the difference of < 0.001 (paired t-test). This represents a 38% reduction from the baseline to the final visit. Table 3 shows mean DED scores at baseline and final visit and mean differences between baseline and final visit after ACE and NCE exposure. Exploratory analyses showed that all seven individual symptoms that constitute the total DED score contribute to the reduction (supplemental figure S1).

**Table 3.** Improvement of DED related symptoms after 4 weeks of thrice daily treatment with I-C eye drops, assessed via the total DED symptom score after ACE and NCE exposure, as primary and secondary endpoint, respectively. Difference is the mean difference in total DED score between baseline (visit 2) and after 4 weeks (visit 4 = final visit) of treatment.

|  | Baseline |  | Visit 4 |  | Mean Difference |  |  |  |
| --- | --- | --- | --- | --- | --- | --- | --- | --- |
|  | n | Mean (SD) | n | Mean (SD) | n | Mean (SD) | 95% CI* | p-value* |
| **ACE** | 28 | 21.89 (11.07) | 28 | 10.00 (8.78) | 28 | -11.89 (8.31) | -15.11; -8.67 | < 0.001 |
| **NCE** | 28 | 21.57 (9.32) | 28 | 13.50 (10.75) | 28 | -8.07 (6.81) | -10.71; -5.43 | < 0.001 |

**Abbreviations:** DED, dry eye disease; n, number of participants in the respective population; SD, Standard Deviation; CI, Confidence Interval; ACE, adverse controlled environment; NCE, normal controlled environment.
\* Paired t-test

Responder rates, ie, the number of patients revealing an improvement of the total DED symptoms at the final visit compared to baseline was analyzed as a key secondary endpoint. The vast majority of participants (26 out of 28 [93%] after ACE and 25 out of 28 [89%] after NCE exposure) recorded a reduction in total DED score between baseline and final visit. Figure 3 shows responder rates for various cut-offs after ACE and NCE exposure.

**Figure 3.**
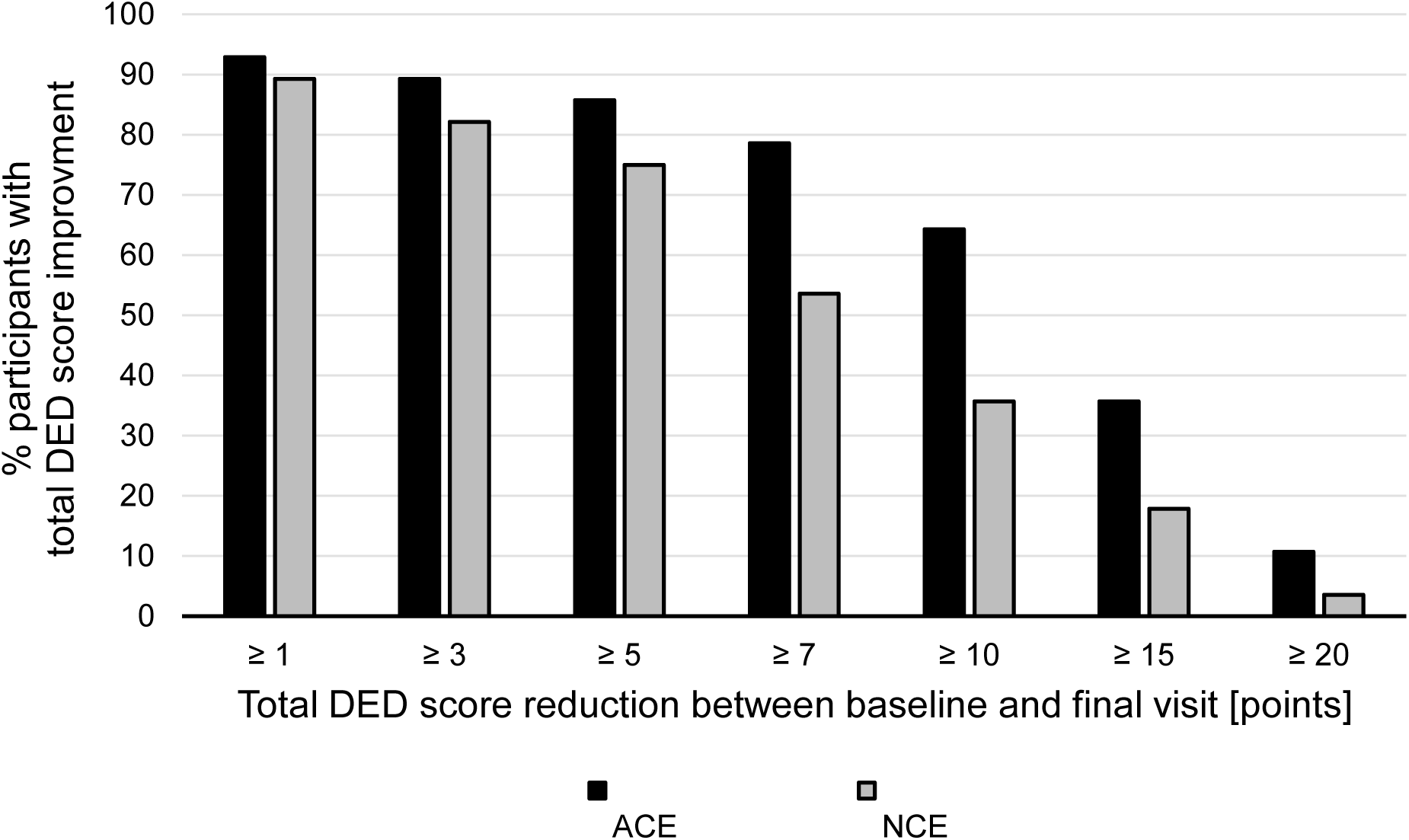
Key secondary endpoint: Responder rates for total DED score. Percentage of participants with improvement of total DED score after 4 weeks of treatment with I-C eye drops, assessed after ACE and NCE exposure. Analysis was carried out in the evaluable FAS, N=28. ACE, adverse controlled environment; DED, dry eye disease; FAS, full analysis set; I-C, iota-carrageenan; NCE, normal controlled environment.

Two additional patient-report outcomes (PROs) were evaluated: the OSDI score and the CDES-Q, both after NCE. The mean (SD) OSDI score declined from 23.80 (5.19) at baseline to 15.81 (9.83) at the final visit, the mean OSDI score difference was −7.75 (95% CI: −10.86; −4.63) with p for the difference between baseline and final visit of < 0.001 (paired t-test). In the CDES-Q, participants were asked at the final visit to assess their change in symptoms as better, same or worse relative to baseline. 21 out of 28 (75%) participants indicated an improvement. Only one participant reported deterioration of the symptoms; the remaining 6 participants reported no difference. Of those who indicated that their symptoms had improved, the improvement ranged from 2 to 9 points. The mean (SD) improvement was 6.10 (2.34) points between final visit and baseline, with a median value of 7 points. Figure 4 shows OSDI score at baseline and final visit (panel A) and CDES-Q results (panel B) after 4 weeks of treatment with I-C eye drops.

**Figure 4.**
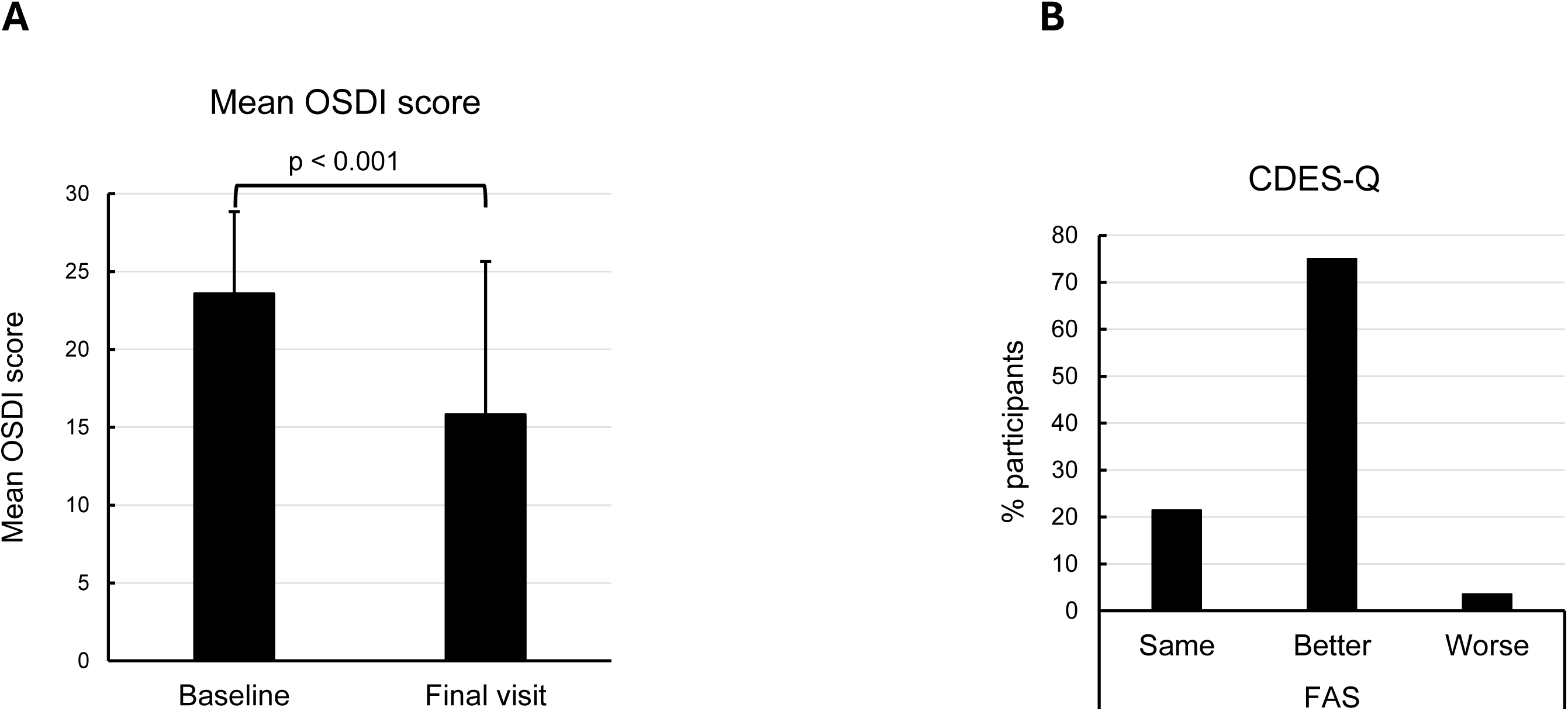
Secondary endpoints: Patient reported outcomes OSDI and CDES-Q. Analysis was carried out in the evaluable FAS, N=28. **A:** Mean OSDI score at baseline and final visit. Error bars represent standard deviation. The p value was derived by paired t-test. **B**: CDES-Q: Proportion of patients at final visit indicating that their eyes feel same, better or worse compared to baseline. OSDI, Ocular Surface Disease Index; CDES-Q, Change in Dry Eye Symptoms Questionnaire; FAS, full analysis set.

Besides patient-reported outcomes, effectiveness was also assessed in terms of corneal and conjunctival integrity. Corneal damage was visualized by fluorescein staining and graded according to the Oxford scale. The majority of graded corneas fell into categories 0 (no damage) and 1 (minimal damage). No participant had grade 4 (marked) or grade 5 (severe) corneal damage, at either visit, both after ACE and NCE exposure. Corneal damage diminished in the course of the clinical investigation, and the reduction was more pronounced after ACE than after NCE. The mean (SD) Oxford score after ACE exposure was 0.89 (0.98) at baseline and 0.52 (0.81) at final visit, and after NCE exposure was 0.66 (0.83) at baseline and 0.50 (0.79) at final visit (Figure 5, panel A). The mean difference between baseline and final visit was −0.38 (95% CI: −0.59; −0.18) after ACE and −0.16 (95% CI: −0.36; 0.02) after NCE exposure, with the p value for the difference between baseline and final visit being < 0.001 after ACE exposure and 0.091 after NCE exposure (confidence interval and p-value obtained by bootstrapping). As expected, exposure to ACE at baseline led to an increase in corneal damage compared to NCE. This increase was no longer observed after four weeks of treatment with I-C eye drops, with p < 0.001 (obtained by bootstrapping) for the difference between baseline and final visit in ACE-induced corneal damage (Figure 5, panel B). Hence, 4 weeks of treatment with I-C drops prevented ACE-induced corneal damage. Count and proportion of participants in each corneal staining category graded according to the Oxford scale based on density of stained dots, at baseline and final visit are shown in supplemental table S1. Grading of corneal staining based on the CCLRU scale yielded similar results (data not shown).

**Figure 5.**
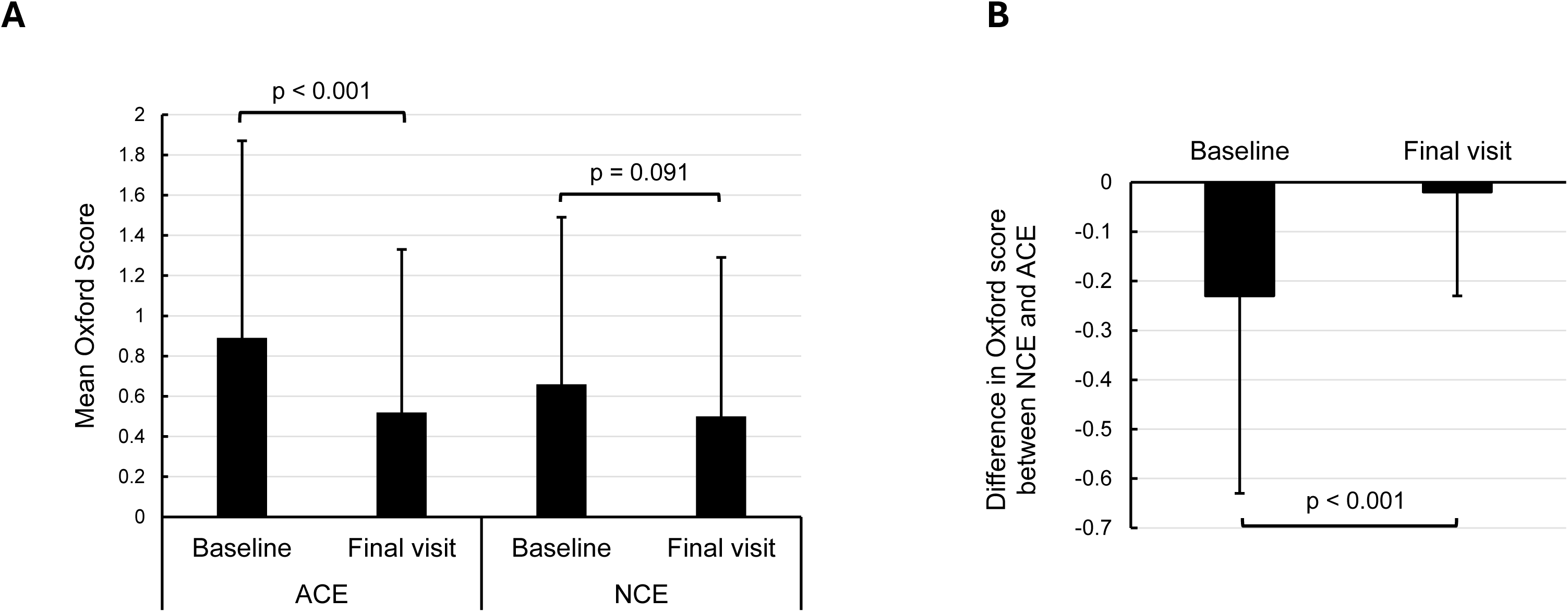
Secondary endpoint: Assessment of ocular surface damage before and after 4 weeks of thrice daily treatment with I-C eye drops, assessed by fluorescein staining of corneal surface damage and rated by the Oxford scale after ACE and NCE exposure. Analysis was carried out in the evaluable FAS; N=28. **A**: Mean Oxford score based on average of right and left eye, at baseline and final visit, after ACE and NCE exposure. Error bars represent standard deviation. P-values for the difference between baseline and final visit were derived by bootstrapping. **B:** Difference in mean Oxford score based on average of right and left eye, between ACE and NCE exposure at baseline and final visit. Error bars represent standard deviation. P-value for the difference between baseline and final visit was derived by bootstrapping. ACE, adverse controlled environment; NCE, normal controlled environment; FAS, full analysis set; I-C, iota-carrageenan.

Conjunctival damage was visualized by lissamine green staining and graded according to the Oxford scale. The majority of graded conjunctivae fell into categories 0 (no damage) and 1 (minimal damage). No participant was assigned to grade 4 (marked) or grade 5 (severe) conjunctival damage, at either visit, both after NCE and ACE exposure. Conjunctival damage diminished in the course of the clinical investigation, and the reduction was more pronounced after ACE than after NCE (supplemental table S2).

In addition to patient-reported outcomes and investigator-assessed ocular surface damage, tear film stability was assessed as an objective measure. Tear film stability was assessed through NIBUT and TBUT. As shown in figure 6 (left part of the diagram), exposure to ACE led to a decreased tear break up time as assessed by TBUT, with a mean (SD) difference in break up time between NCE and ACE of 0.83 (1.10) seconds. After the 4 weeks of treatment with I-C eye drops, the tear film destabilizing effect of ACE exposure was no longer observed, with mean (SD) difference in TBUT between NCE and ACE at the final visit of only 0.09 (1.29) seconds. These results demonstrate the tear film-stabilizing effect of I-C eye drops. Similar results were obtained by NIBUT (figure 6, right part of the diagram). The investigational treatment showed no effect on tear film evaporation, nor on tear production according to the Schirmer test.

**Figure 6.**
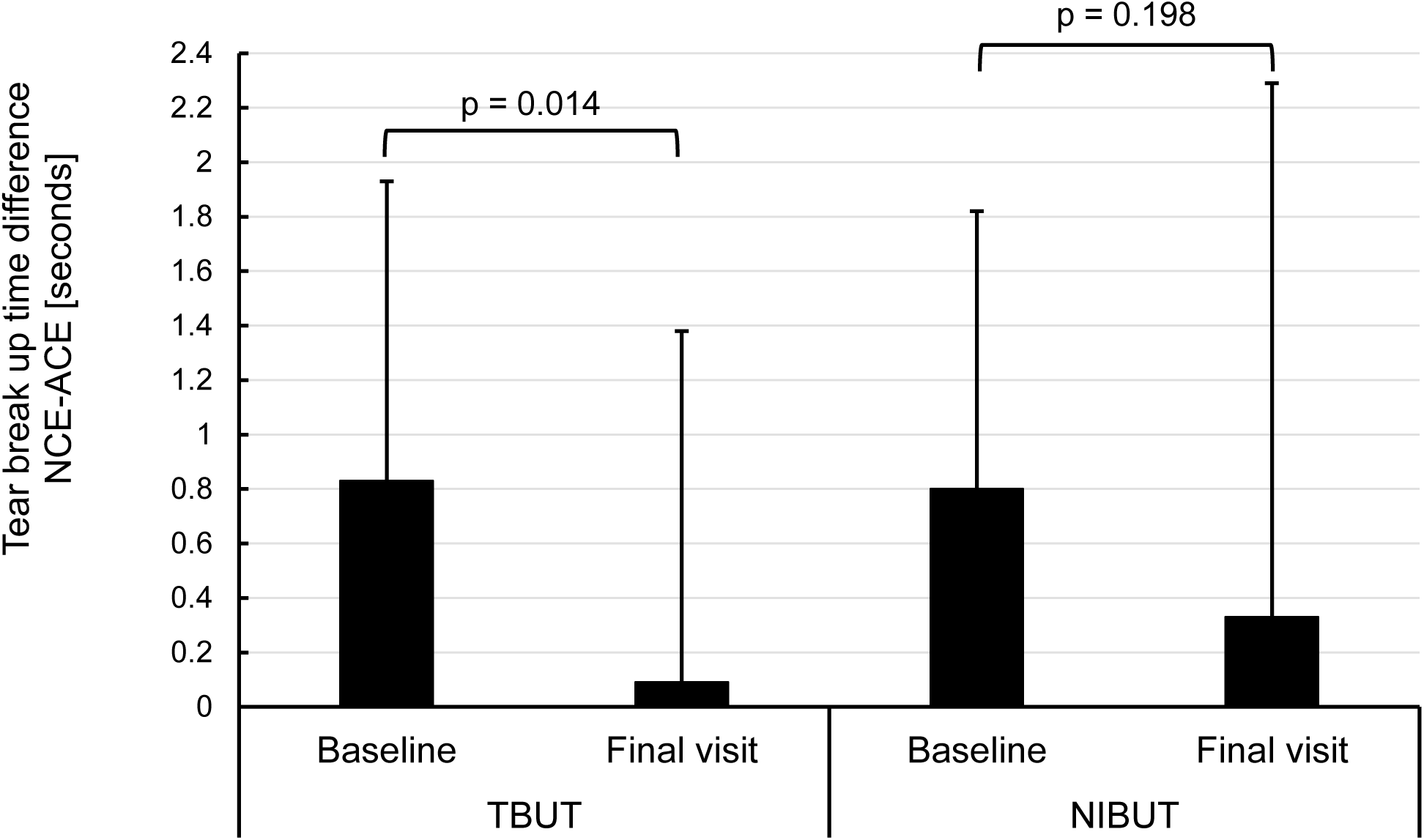
Secondary endpoint: Tear film stability assessed through fluorescein-based (TBUT) and non-invasive (NIBUT) tear film break-up time measurement. Difference between NCE and ACE exposure at baseline and final visit, evaluated by TBUT and NIBUT. Error bars represent standard deviation. P-values for the difference between baseline and final visit in ACE-induced break-up time were derived by bootstrapping. Analysis was carried out in the evaluable FAS; N=28. NIBUT, non-invasive tear breakup time; TBUT, tear film breakup time; ACE, adverse controlled environment; NCE, normal controlled environment; FAS, full analysis set.

### Safety and Tolerability

The tolerability of the IP was assessed by participants by means of a Likert-type scale at baseline and final visit (Figure 7). The vast majority of participants (90%) rated tolerability as good or very good already after the first application. One participant rated tolerability as neutral, two as bad, and none as very bad. After four weeks, tolerability ratings had further improved, with 93% of the participants rating tolerability as good or very good, and two as neutral. Importantly, no participant reported poor tolerability after the four weeks treatment with I-C eye drops.

**Figure 7.**
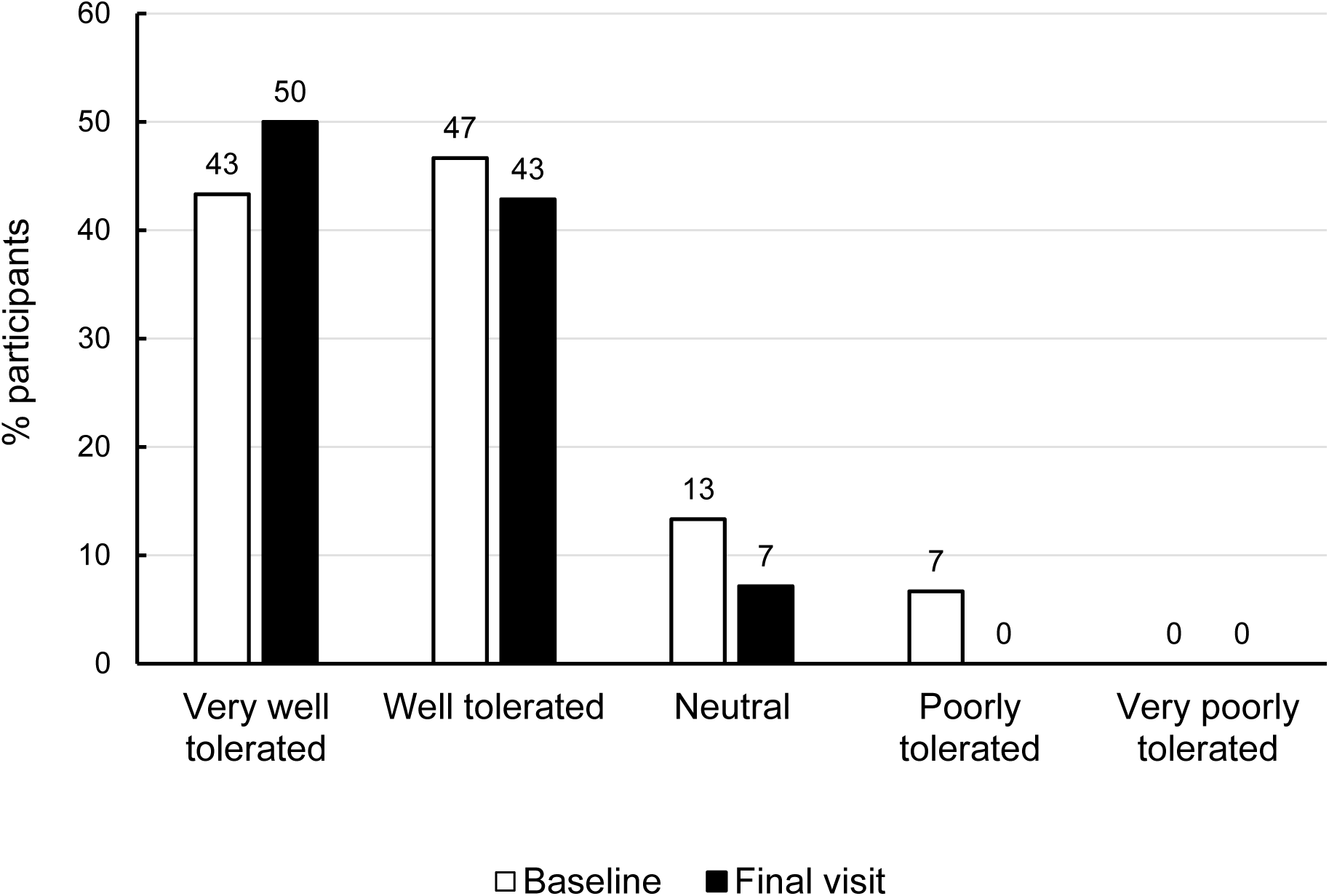
Participant-reported tolerability of iota-carrageenan (I-C) eye drops on a Likert-type scale. Analysis was carried out in the safety population; N=30 at baseline and N=28 at final visit.

Table 4 summarizes adverse events (AEs) reported in this clinical investigation. 21 out of 30 (70%) participants in the safety population reported a total of 107 AEs. All AEs concerned the eyes, with 20 participants with system organ class (SOC) eye disorders, and one with SOC infection/infestation (hordeolum). The AEs that affected most participants were eye pain (reported by 10/30 [33%] participants), eye pruritus and blurred vision (each reported by 8/30 [27%] participants (supplemental table S3). All reported AEs were transient and mild. One AE (hordeolum) was resolving, all other AEs had already resolved at the respective participant’s last visit. 97 out of 107 AEs were considered possibly related to the IP, but none was considered definitely related (supplemental table S3). Two participants terminated their participation in the clinical investigation early due to adverse events: One due to hordeolum, which was not related to the IP, and one due to eye pruritus and foreign body sensation which were possibly related to the IP. None of the AEs concerning participants included in the effectiveness analysis required medication. No deaths, serious adverse events or other significant adverse events occurred during the clinical investigation. No device deficiencies or unanticipated adverse device effects have been reported.

**Table 4.** Adverse events recorded throughout the clinical investigation in the safety population.

| Participants with | n (%) |
| --- | --- |
| At least one AE | 21 (70.00) |
| At least one related AE, mild (grade 1) | 21 (70.00) |
| At least one related AE, moderate (grade 2) | 0 |
| At least one related AE, severe (grade 3) | 0 |
| At least one serious AE, any | 0 |
| At least one AE leading to death | 0 |
| At least one AE leading to early termination | 2 (6.67) |
| At least one related AE leading to early termination | 1 (3.33) |

**Abbreviations:** n = number of subjects with adverse event; AE, adverse event.
Note: Participants are counted once for each category regardless of the number of events.

Visual acuity showed only minor fluctuations over the course of the clinical investigation (data not shown). Slit lamp examinations of the ocular surface and anterior intraocular structures analyzed as safety parameters included evaluation of conjunctival redness, blepharitis, meibomian gland dysfunction, anterior segment inflammation and corneal findings. A tendency towards reduction of conjunctival redness over the 4-weeks treatment course was observed after ACE exposure. No changes between baseline and final visits were observed for blepharitis and meibomian gland dysfunction. No anterior segment inflammation or corneal findings were recorded in any participant at any visit.

Most recorded IOP values were in the normal range; only two participants had IOP values slightly above the normal range (22-23 mmHg), and three participants had IOP values slightly below the normal range (7-9 mmHg). Importantly, no IOP value was considered pathologic by the investigator, and no participant received medical treatment due to elevated or reduced IOP during the clinical investigation.

No adverse effects on any of the assessed safety parameters were observed.

## Discussion

DED affects people throughout the world and is one of the most frequent causes of patients to visit eye care practitioners.^24^ There is a wide variety of lubricating ophthalmic solutions, gels and ointments on the market. However, there are still significant unmet needs, such as tolerability issues, limited ability to promote regeneration or healing of ocular tissue damage, or delay of several weeks or months in the onset of therapeutic effect, for the adequate management of this disease.^25^

In this article we present results of a non-randomized, uncontrolled, open-label, single arm, monocentric clinical investigation aimed to evaluate the effectiveness and safety of I-C eye drops for the treatment of DED. Participants suffering from mild-to-moderate DED were treated with I-C eye drops thrice daily for 28 ± 4 days. The effectiveness of I-C eye drops was assessed in the evaluable FAS of 28 participants, which included all participants to whom the IP has been administered and who had data available for both baseline and final visit. Effectiveness evaluation was based on patient-reported outcomes, as well as on investigator-rated outcomes before and at the end of the treatment period. To test the ability of I-C eye drops to protect against the development of ocular surface damage and symptom induction/exacerbation triggered by adverse environmental conditions, effectiveness was rated after exposure of participants to NCE and to ACE with airflow and reduced humidity.

Our results show that after 4 weeks of three times daily treatment with I-C eye drops, DED symptoms significantly decreased compared to baseline. DED symptom severity as rated by participants on numerical rating scales was reduced by 54% after ACE exposure, and by 37% after NCE exposure. These results were backed by a reduction of the OSDI by 7.75 points. The minimum clinically important difference in OSDI is in the range of 4.5 to 7.3 points for mild to moderate DED.^26^ The observed OSDI reduction can thus be considered a clinically meaningful improvement. Interestingly, all rated DED symptoms that are subjectively perceptible by patients (foreign body sensation, burning/stinging, itching, pain, sticky feeling, blurred vision, and photophobia), and all three sections of the OSDI contributed to the improvement, showing that I-C eye drops benefit all aspects of DED and can therefore provide relief to a large variety of patients with different manifestations of DED.

In addition to patient-reported outcomes, effectiveness was confirmed by investigator-rated surface damage of the cornea and conjunctiva. Corneal fluorescein staining, quantified based on the Oxford grading scale, reflects corneal epithelium integrity and is therefore an important diagnostic tool for corneal damage.^23^ The difference in Oxford score between NCE and ACE at baseline demonstrates the capacity of the selected ACE conditions to elicit ocular surface damage. However, this difference was reduced or nearly abolished after four weeks of three times daily treatment with I-C eye drops. These results demonstrate that I-C eye drops not only provided effective symptom relief but also prevented formation of ocular surface damage induced by adverse environmental conditions such as low humidity and airflow, demonstrating pronounced lubrication properties of I-C eye drops.

Impaired tear film stability is fundamental for the diagnosis of DED and is routinely assessed in clinical practice by measuring TBUT.^23^ The marked difference in tear film stability between NCE and ACE recorded at baseline indicates the harmful effect of the experimental conditions on tear film stability. However, analogous to our observations on ocular surface damage, this difference had been reduced or nearly abolished after four weeks of thrice daily treatment with I-C eye drops. The increase in TBUT from baseline to final visit indicated improved corneal wettability and highlight the water-retaining, lubricating and thus tear film stabilizing properties of I-C eye drops, similar to what has previously been demonstrated for hyaluronic acid.^27–31^

Effectiveness of I-C eyedrops was paired with remarkable tolerability, with 90% of participants rating tolerability as good or very good already after the first application and increased further to 93% after 4 weeks of treatment. No participant rated tolerability as poor, and only 2 participants rated it as neutral. This is not commonly observed in clinical studies of comparable products and exceed the acceptance rate of many HA-based formulations.^32–34^

The high degree of subjectively rated excellent tolerability is paired with reassuring results of safety-related ophthalmic measurements of visual acuity, intraocular pressure and slit-lamp evaluation of the morphology and anatomy of the eye surface and adnexa.

In sum, these results demonstrate that I-C eye drops significantly alleviated signs and symptoms of mild-to-moderate DED and were safe and exceptionally well tolerated.

Mechanistically, these effects likely stem from the physicochemical properties of I-C, a long-chain high molecular weight polysaccharide containing highly polar groups. The thickening and emulsifying properties of carrageenan have long been known and exploited in the food and the cosmetics industry. The excellent water-holding, thickening, gel building and mucoadhesive properties of carrageenan are also exploited in pharmaceutical applications for various forms of drug delivery,^36^ and carrageenan films have proven useful as mucoadhesive in ocular drug delivery systems.^13–15,37^ From a mechanistic perspective, we therefore hypothesize that these properties together contribute to the observed beneficial effect: I-C forms a mucoadhesive film on the ocular surface, it enhances viscosity of the tear film, improves surface retention, and counteracts dehydration.

For eye drops to be effective in alleviating DED symptoms, it is essential to strike a balance of viscosity. A higher degree of viscosity provides a longer duration of effect due to extended ocular residence, and is therefore associated with greater improvements in signs and symptoms of DED compared with standard low-viscosity lubricating eye drops.^35–37^ If eye drops are too viscous, on the other hand, this may reduce tolerability due to blur, stickiness, and build-up of residue on the lids and lashes, which limits their application.^38,39^ Blurred vision is frequently observed in clinical studies of HA-based ophthalmic products.^13,40^ In our clinical investigation, patient-reported stickiness and blurred vision improved by approximately 60% after four weeks of treatment. We hence assume that, based on the characteristics of I-C and our selected formulation, I-C eye drops confer the optimal degree of viscosity, balancing contact time, prolonged lubrication and relief from dryness with minimal shear-stress and build-up of residue.

A key advantage of I-C eye drops is the clinically meaningful effectiveness after a relatively short treatment duration of four weeks, with low application frequency of three times a day.

Comparable products based on HA or CMC are applied up to 6-8 times per day or sometimes even ad libitum without upper limit of daily applications, for up to 3 months.^32,33,41^ In should be noted that in our clinical investigation participants with mild-to-moderate DED (OSDI score ≥ 13 and < 33 points) were included, while the cited studies on HA- or CMC-based eye drops were conducted in moderate to severe DED patients (with an OSDI score of ≥18,^33^ ≥22,^32,33^ or ≥28^41^). Unlike other substances used for DED treatment, like the highly controversial “forever”-chemical perfluorohexyloctane,^7,42^ I-C is a natural, well-studied substance. No health hazards have been documented for I-C so far. The toxicological aspects of carrageenans have been thoroughly evaluated and have been established to have minimal or no adverse physiological effects.^43^ I-C has been designated as “Generally Recognized As Safe” (GRAS status) by the FDA for food and for topical applications.^44^

Additionally, I-C eye drops are a preservative-free formulation, as due to significant toxic effects induced by preservatives ^45^, the use of preservative-free eye drops is particularly recommended to patients requiring frequent instillation of eye drops.^46^ Hence, I-C eye drops may offer a decisive advantage over preserved competitor products.

Limitations of the clinical investigation include the open-label, single arm design that may allow introduction of bias, however this type of design has been used by others and is generally deemed fit-for-purpose.^47,48^ The evaluated objective clinical signs, which are less prone to expectation bias, confirm treatment effectiveness after the 4-weeks treatment course. An additional limitation is the treatment duration of four weeks, which allows only limited conclusions on long-term effectiveness, tolerability and safety.

Strengths of the investigation include the multidimensionality of the total DED score, used as primary outcome measure capturing all relevant patient-perceived DED symptoms of discomfort and visual disturbance, as recommended by the TFOS DEWS III Report.^49^ This subjective measures reflect significant patients’ quality of life (QOL), which is considered an important determinant for DED treatment effectiveness.^50^ Assessment of QOL in the form of patient-reportd outcomes is regarded a standard outcome measure in clinical trials, acknowledging the fact that objective clinical signs often correlate poorly with symptoms reported by patients and their impact on QOL.^51–53^ Objective signs like corneal staining are late findings in the development of disease and are variable over time owing to the instability of the tear film, a hallmark of DED.^54,55^ Moreover, especially in early or mild DED, the presence of hyperalgesia can cause significant ocular discomfort without any signs of tissue damage.^56,57^ Therefore, the collection of qualitative patient-reported outcome data is considered essential for comprehensive evaluation and optimal treatment management. In addition to the total DED score, which has been developed and applied only recently,^19^ two additional, validated questionnaires were used to test and confirm the robustness of findings: the OSDI and the CDES-Q. The OSDI is the most widely used patient questionnaire for DED in clinical studies, with > 600 records).^58^ The CDES-Q is a questionnaire used to detect changes from baseline.^59^ The treatment effect was confirmed with all used questionnaires. In addition, patient-reported results were supported by investigator-rated and objective findings, confirming robustness and reliability of the effect.

An additional strength is the DED assessment after NCE and ACE. Standardized ACE exposure induced DED symptoms and corneal and conjunctival surface damage and destabilized the tear film. This shows the relevance of the tested conditions and enabled the discovery of significant effects with the limited available sample size and a population with predominantly mild DED, in which objective signs are often not yet manifest to a measurable degree under basal conditions.^60^ The extent to which these findings can be generalized to other populations and clinical contexts requires further investigation. Based on the results presented here, we expect a similar or even more pronounced effect in patients of severe DED with an elevated baseline level of signs and symptoms. We further believe that based on the low application frequency (1 drop per eye, thrice daily), I-C eye drops are well suited as supplementary therapy in addition to baseline medication for patients who experience DED as symptom of an underlying disease such as Sjögren’s syndrome.

## Conclusion

In conclusion, I-C eye drops hydrate and lubricate the ocular surface; they contribute to repair (or prevent further formation/accumulation) of ocular surface damage, and stabilize the tear film, leading to significantly reduced ocular irritation resulting from dryness. Thus, I-C eye drops present a well tolerable, safe, preservative-free treatment option for DED. Effectiveness is achieved for the entire symptom spectrum and therefore suited to a large variety of disease manifestations.

### Ethics Approval and Informed Consent

The clinical investigation was conducted in Spain in accordance with the Declaration of Helsinki on Ethical Principles for Medical Research Involving Human Subjects, the International Council for Harmonisation Guideline on Good Clinical Practice, and all applicable local regulatory requirements and laws. The clinical investigation was approved by the responsible Ethics Committee (Comité de Ética de la Investigación con Medicamentos Área de Salud Valladolid Este) at the meeting of 23-11-2023 (documented in meeting minutes number 23 of 2023). Informed consent was obtained from all clinical investigation participants.

## Disclosure

NUM, HD, and EPG are employees of Marinomed Biotech AG. EPG reports grants from FFG outside the submitted work. EPG and SRN are holders of a relevant patent: EP24167732. MS has received consulting fees from Marinomed Biotech AG. The other authors have no competing interests in this work.

After completion of the clinical investigation, Marinomed’s iota-carrageenan product portfolio, including the iota-carrageenan eyedrops, has transitioned to Unither Pharmaceuticals (Paris, France).

**Supplemental Figure S1** Mean difference in individual DED symptom scores between baseline and final visit, as recorded after ACE (A) and NCE (B) exposure. Analysis was carried out in the evaluable FAS; N=28. Error bars represent 95% confidence intervals.

ACE, adverse controlled environment; DED, dry eye disease; FAS, full analysis set; NCE, normal controlled environment.

## Data Availability

All data produced in the present study are available upon reasonable request to the authors.

