## Supplemental Figure S1 for "Safety and Effectiveness of Iota-Carrageenan Eye Drops for the Treatment of Dry Eye Disease"

**Supplemental figure S1**: Mean difference in individual DED symptom scores between baseline and final visit, as recorded after ACE (A) and NCE (B) exposure. Analysis was carried out in the evaluable FAS; N=28. Error bars represent 95% confidence intervals. ACE, adverse controlled environment; DED, dry eye disease; FAS, full analysis set; NCE, normal controlled environment.

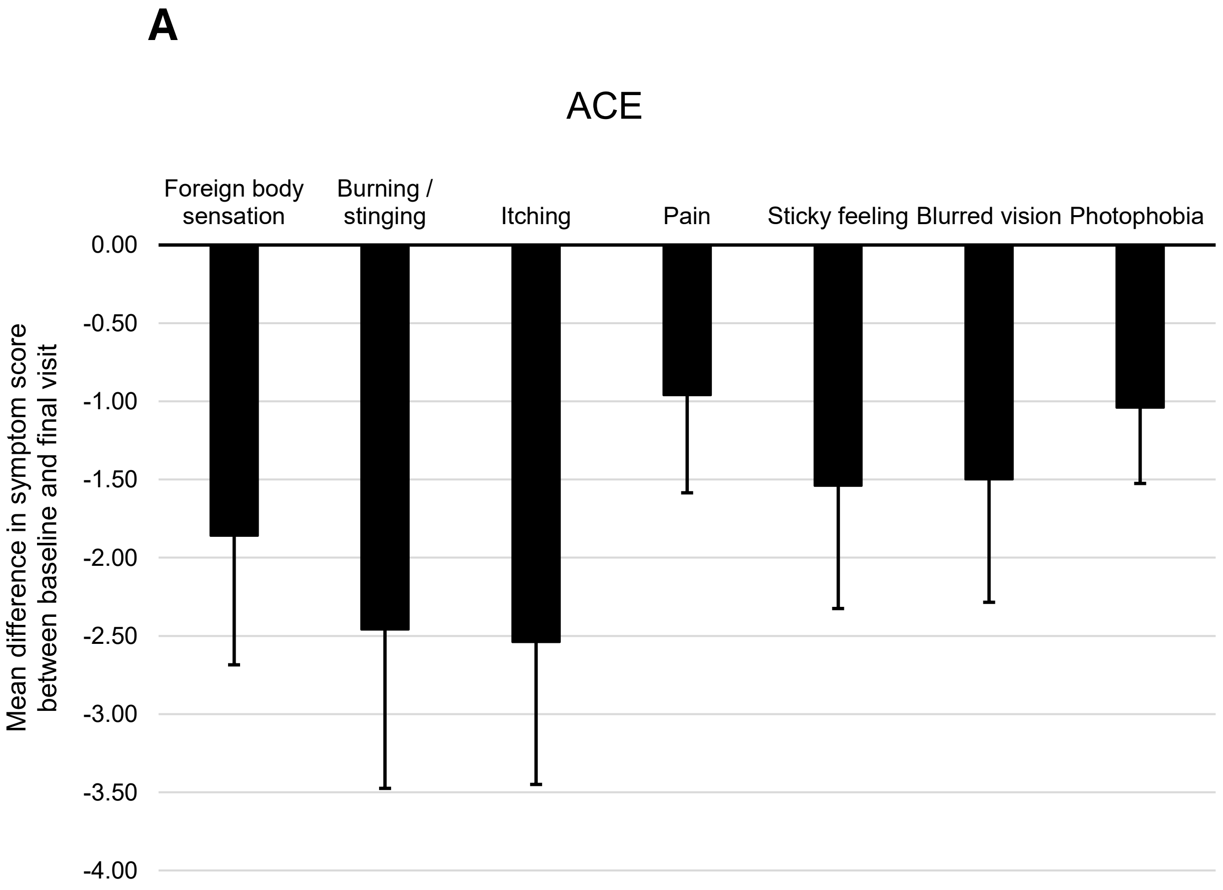


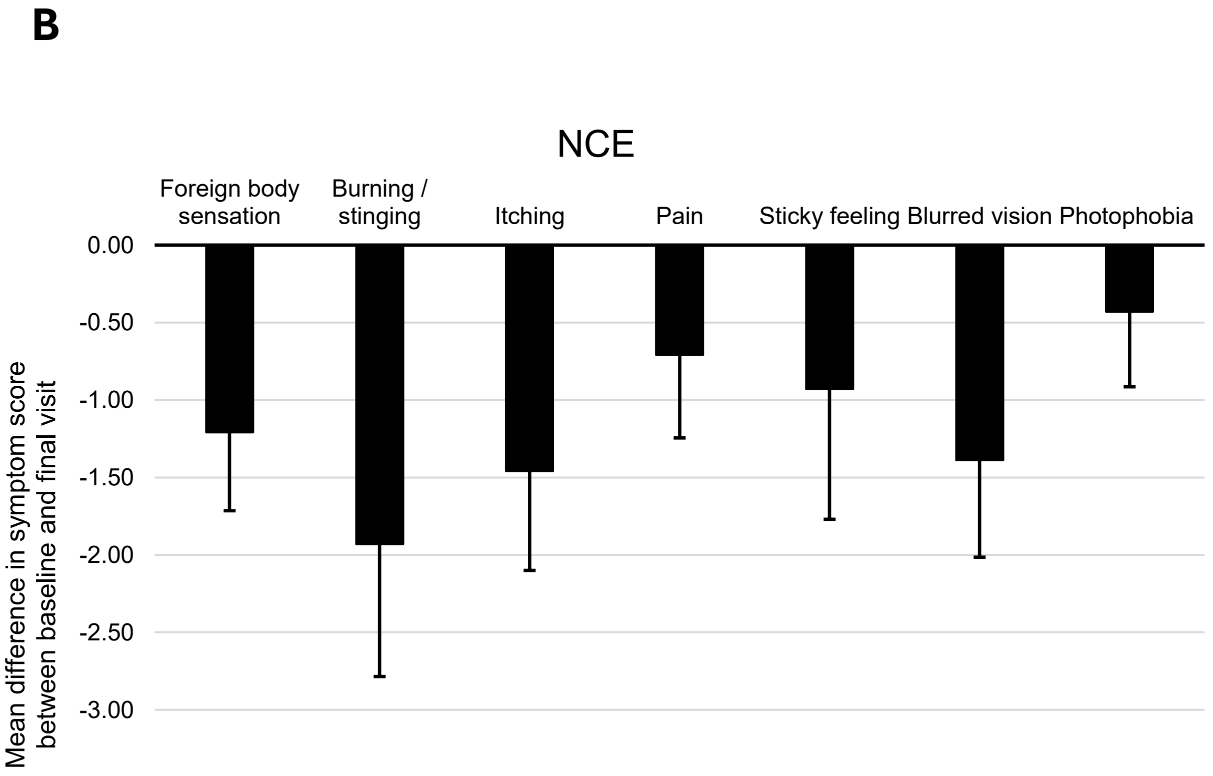
