## Supplemental Tables for "Safety and Effectiveness of Iota-Carrageenan Eye Drops for the Treatment of Dry Eye Disease"

Clinical Trial Report

Nicole Unger-Manhart et al.

### Supplemental Tables

**Table S1** Distribution of participants across corneal staining categories based on density of stained dots graded according to the Oxford scale at baseline (visit 2) and final visit (visit 4), for the right and left eye, for the evaluable full analysis set after exposure to normal controlled environment (NCE) and to adverse controlled environment (ACE).

|  | **ACE** | | | | **NCE** | | | |
| --- | --- | --- | --- | --- | --- | --- | --- | --- |
|  | **OD Visit 2** | **OS Visit 2** | **OD Visit 4** | **OS Visit 4** | **OD Visit 2** | **OS Visit 2** | **OD Visit 4** | **OS Visit 4** |
|  | **n (%)** | **n (%)** | **n (%)** | **n (%)** | **n (%)** | **n (%)** | **n (%)** | **n (%)** |
| Grade 0 (absent) | 12 (42.86) | 13 (46.43) | 19 (67.86) | 19 (67.86) | 12 (42.86) | 18 (64.29) | 18 (64.29) | 20 (71.43) |
| Grade 1 (minimal) | 9 (32.14) | 9 (32.14) | 4 (14.29) | 5 (17.86) | 13 (46.43) | 6 (21.43) | 6 (21.43) | 4 (14.29) |
| Grade 2 (mild) | 3 (10.71) | 4 (14.29) | 4 (14.29) | 3 (10.71) | 1 (3.57) | 2 (7.14) | 3 (10.71) | 3 (10.71) |
| Grade 3 (moderate) | 4 (14.29) | 2 (7.14) | 1 (3.57) | 1 (3.57) | 2 (7.14) | 2 (7.14) | 1 (3.57) | 1 (3.57) |
| Grade 4 (marked) | 0 | 0 | 0 | 0 | 0 | 0 | 0 | 0 |
| Grade 5 (severe) | 0 | 0 | 0 | 0 | 0 | 0 | 0 | 0 |

**Abbreviations** OD, Oculus dexter; OS, Oculus sinister; n, number of participants with specified outcome.

**Table S2** Distribution of participants across conjunctival staining categories based on density of stained dots graded according to the Oxford scale at baseline (visit 2) and final visit (visit 4), for the right and left eye, for the evaluable full analysis set after exposure to normal controlled environment (NCE) and to adverse controlled environment (ACE). Nasal (upper panel) and temporal conjunctiva (lower panel) were analyzed separately.

**Nasal conjunctiva:**

|  | **ACE** | | | | **NCE** | | | |
| --- | --- | --- | --- | --- | --- | --- | --- | --- |
|  | **OD Visit 2** | **OS Visit 2** | **OD Visit 4** | **OS Visit 4** | **OD Visit 2** | **OS Visit 2** | **OD Visit 4** | **OS Visit 4** |
|  | **n (%)** | **n (%)** | **n (%)** | **n (%)** | **n (%)** | **n (%)** | **n (%)** | **n (%)** |
| Grade 0 (absent) | 15 (53.57) | 9 (32.14) | 16 (57.14) | 16 (57.14) | 15 (53.57) | 11 (39.29) | 16 (57.14) | 16 (57.14) |
| Grade 1 (minimal) | 7 (25.00) | 16 (57.14) | 7 (25.00) | 10 (35.71) | 9 (32.14) | 14 (50.00) | 8 (28.57) | 10 (35.71) |
| Grade 2 (mild) | 5 (17.86) | 1 (3.57) | 5 (17.86) | 2 (7.14) | 4 (14.29) | 3 (10.71) | 3 (10.71) | 1 (3.57) |
| Grade 3 (moderate) | 1 (3.57) | 2 (7.14) | 0 | 0 | 0 | 0 | 1 (3.57) | 1 (3.57) |
| Grade 4 (marked) | 0 | 0 | 0 | 0 | 0 | 0 | 0 | 0 |
| Grade 5 (severe) | 0 | 0 | 0 | 0 | 0 | 0 | 0 | 0 |

**Temporal conjunctiva:**

|  | **ACE** | | | | **NCE** | | | |
| --- | --- | --- | --- | --- | --- | --- | --- | --- |
|  | **OD Visit 2** | **OS Visit 2** | **OD Visit 4** | **OS Visit 4** | **OD Visit 2** | **OS Visit 2** | **OD Visit 4** | **OS Visit 4** |
|  | **n (%)** | **n (%)** | **n (%)** | **n (%)** | **n (%)** | **n (%)** | **n (%)** | **n (%)** |
| Grade 0 (absent) | 18 (64.29) | 20 (71.43) | 24 (85.71) | 23 (82.14) | 17 (60.71) | 19 (67.86) | 23 (82.14) | 24 (85.71) |
| Grade 1 (minimal) | 9 (32.14) | 7 (25.00) | 4 (14.29) | 5 (17.86) | 11 (39.29) | 7 (25.00) | 5 (17.86) | 4 (14.29) |
| Grade 2 (mild) | 1 (3.57) | 1 (3.57) | 0 | 0 | 0 | 2 (7.14) | 0 | 0 |
| Grade 3 (moderate) | 0 | 0 | 0 | 0 | 0 | 0 | 0 | 0 |
| Grade 4 (marked) | 0 | 0 | 0 | 0 | 0 | 0 | 0 | 0 |
| Grade 5 (severe) | 0 | 0 | 0 | 0 | 0 | 0 | 0 | 0 |

**Abbreviations** OD, Oculus dexter; OS, Oculus sinister; n, number of participants with specified outcome.

**Table S3** Adverse Events on participant level by MedDRA system organ class and preferred term by causal relationship to the investigational product, on participant level. Analysis was carried out in the participant population.

| **MedDRA SOC** | **MedDRA PT** | **No Relationship** | | | **Any Relationship** | | | **Total (N=30)** |
| --- | --- | --- | --- | --- | --- | --- | --- | --- |
|  |  | **Unrelated** | **Unlikely related** | **Total no relationship** | **Possibly related** | **Related** | **Total any relationship** |  |
|  |  | n (%) | n (%) | n (%) | n (%) | n (%) | n (%) | n (%) |
| Participants with any SOC | Participants with any PT | 1 (3.33) | 4 (13.33) | 5 (16.67) | 20 (66.67) | 0 | 20 (66.67) | 21 (70.00) |
| Infections and infestations | Hordeolum | 1 (3.33) | 0 | 1 (3.33) | 0 | 0 | 0 | 1 (3.33) |
| Eye disorders | Abnormal sensation in eye | 0 | 0 | 0 | 3 (10.00) | 0 | 3 (10.00) | 3 (10.00) |
|  | Dry eye | 0 | 0 | 0 | 1 (3.33) | 0 | 1 (3.33) | 1 (3.33) |
|  | Eye discharge | 0 | 0 | 0 | 4 (13.33) | 0 | 4 (13.33) | 4 (13.33) |
|  | Eye irritation | 0 | 0 | 0 | 4 (13.33) | 0 | 4 (13.33) | 4 (13.33) |
|  | Eye pain | 0 | 1 (3.33) | 1 (3.33) | 10 (33.33) | 0 | 10 (33.33) | 10 (33.33) |
|  | Eye pruritus | 0 | 0 | 0 | 8 (26.67) | 0 | 8 (26.67) | 8 (26.67) |
|  | Eyelids pruritus | 0 | 0 | 0 | 1 (3.33) | 0 | 1 (3.33) | 1 (3.33) |
|  | Foreign body sensation in eyes | 0 | 1 (3.33) | 1 (3.33) | 3 (10.00) | 0 | 3 (10.00) | 3 (10.00) |
|  | Lacrimation increased | 0 | 2 (6.67) | 2 (6.67) | 1 (3.33) | 0 | 1 (3.33) | 3 (10.00) |
|  | Ocular hyperaemia | 0 | 0 | 0 | 1 (3.33) | 0 | 1 (3.33) | 1 (3.33) |
|  | Blurred vision | 0 | 0 | 0 | 8 (26.67) | 0 | 8 (26.67) | 8 (26.67) |

**Abbreviations** MedDRA, Medical Dictionary for Regulatory Activities; SOC, system organ class; PT, preferred term; n, number of participants with AE.
