## Supplemental Methods for "Safety and Effectiveness of Iota-Carrageenan Eye Drops for the Treatment of Dry Eye Disease"

### **Numerical Rating Scales (NRS) on Dry Eye Disease (DED) Symptoms**

The primary endpoint of this study was a composite score on ocular symptoms related to DED, including a) sensations of foreign body, b) burning or stinging, c) itching, pain, d) sticky feeling, e) blurred vision, and f) photophobia, each evaluated on 0-10 NRS, obtained by simple summation of items (total score range: 0-70 points).

### **Ocular Surface Disease Index (OSDI)**

This tool was developed about 25 years ago to quickly assess the symptoms of ocular irritation in DED and how they affect functioning related to vision.^1^ It features 12 items and is scored with respect to the status along the last week. These produce three subscales: ocular symptoms, vision-related function, and environmental triggers. Patients rate their responses on 0 to 4 scale. A final score is calculated which ranges from 0 to 100 with scores ≤12 representing normal, >12 to ≤22 representing mild DED, >22 to ≤32 representing moderate DED, and >32 representing severe DED.^1,2^

### **Change in Dry Eye Symptoms Questionnaire (CDES-Q)**

This is a questionnaire recently developed to detect changes in DED symptoms, since DED tools usually provide only indirect measures of change.^3^ Although not composite per se, this tool provides a comprehensive effect measure since it gathers a global impression of change, which has been shown to portend rich and useful information in other conditions, like chronic neuropathic pain, in which subjectivity poses considerable difficulties when doing clinical research.^4^ It consists of two questions, one asks for the change in symptoms relative to a pre-defined recalling period, and the other quantifies such change on a 0 to 10 visual graded scale.^3^

### **Best Corrected Visual Acuity**

Visual acuity (VA) was assessed as per the investigators’ routine practice using optimum refractive error correction as recorded in patient’s medical history. A screen with 100% illumination and a suitable Early Treatment of Diabetic Retinopathy Study (ETDRS) charts placed at 4 m in front of the patient were used. The overhead room illumination was switched on during VA testing. The patient was asked to begin reading the largest line of the optotype he/she can easily read, and proceed further down until three or more letters are missed on a given line. VA measurements were recorded as logMAR scores, for which each letter has a value of 0.02 log units.

### **Evaluation of Tear Film Stability**

Tear film stability was assessed through tear film breakup time (TBUT) and non-invasive tear breakup time (NIBUT) as well as through tear evaporation rate. TBUT was conducted using cobalt blue light in a slit lamp and a Wratten #12 yellow filter (Eastman Kodak, Rochester, NY, USA) after instilling sodium fluorescein through impregnated strips with saline solution. NIBUT was measured using the Easy Tear® VIEW+ Tearscope (EASYTEAR S.R.L., Trento, Italy). Patients were instructed to blink naturally three times and then to cease blinking until instructed. Both (TBUT and NIBUT) were defined as the interval of time elapsed between a complete blink and the appearance of the first break in the tear film.^5^ Three measurements were conducted and the average was calculated.

Tear evaporation rate was measured using the Eye Vapometer (Delfin Technologies Ltd. Kuopio, Finlandia). First, the patient was asked to blink normally and remain in the primary gaze position while three measurements were taken. Then, they were asked to close their eyes while three more measurements were taken. The final result is obtained from the difference in tear evaporation between the average evaporation with eyes open and eyes closed in order to take into account evaporation from the skin of the eyelids and surrounding tissue.

### **Evaluation of Damage to Ocular Surface**

The cornea and conjunctiva were assessed after instilling sodium fluorescein and lissamine green, respectively, through impregnated strips with saline solution using the slit lamp. The Oxford grading scale was used for the evaluation of cornea and nasal and temporal conjunctiva staining. This scale ranges from 0 to 5: Grade 0 (absent staining), Grade 1 (minimal staining), Grade 2 (mild staining), Grade 3 (moderate staining), Grade 4 (marked staining) and Grade 5 (severe staining).^6^ In addition, the type (micropunctate, macropunctate, coalescent macopunctate and patch) and extent (1-15%, 16-30%, 31-45%, >45%) of corneal staining were assessed with the Cornea and Contact Lens Research Unit (CCLRU) scale.^7^

### **Evaluation of Inflammation of Ocular Surface**

Conjunctival redness was assessed by a slit lamp using the Efron scale, which classifies it into Grade 0 (normal), Grade 1 (trace), Grade 2 (mild), Grade 3 (moderate) and Grade 4 (severe).^8^ The present or absence of corneal findings and anterior segment inflammation were also evaluated by a slit lamp.

### **Evaluation of Eyelid Aspects**

Blepharitis and Meibomian gland dysfunction were assessed using a slit lamp and the Efron scale as Grade 0 (normal), Grade 1 (trace), Grade 2 (mild), Grade 3 (moderate) and Grade 4 (severe).

### **Evaluation of Tear Volume**

Tear production was assessed through the Schirmer test without anesthesia. The folded end of a Schirmer paper strip was hooked over the temporal one-third of the lower lid margin and kept for 5 minutes while the eyes were closed. The score was expressed as the length of wetting of the paper strip.

### **Evaluation of Intraocular Pressure**

Intraocular pressure was measured with the iCare IC100 non-contact tonometer (Icare Finland Oy, Vantaa, Finland).
